# Analyzing Conspiratorial Content Across Singapore-Based Telegram Groups

**DOI:** 10.1101/2025.07.15.25331450

**Authors:** Abhay Goyal, Val Alvern Cueco Ligo, Lam Yin Cheung, Roy Ka-Wei Lee, Koustuv Saha, Edson C. Tandoc, Navin Kumar

**Affiliations:** Nimblemind, USA; Independent Researcher, USA; Singapore University of Technology and Design, SG; University of Illinois Urbana-Champaign, USA; Nanyang Technical University, SG

**Keywords:** Telegram, Information flow, Singapore, Misinformation

## Abstract

Telegram has emerged as a key platform for the circulation of conspiratorial narratives. We examine conspiratorial discourse within Singapore-based Telegram groups from 2021–2025. We analyze over 10 million words from three Telegram groups. We developed a logistic regression classifier to detect conspiratorial content, achieving an F1 score of 0.74 and expert-validated labeling accuracy of 72%. Topic models indicated dominant themes centered around elite control, vaccine risks, and globalist agendas. While most users rarely posted conspiratorial content, a small, highly active minority accounted for most of such messages. These users frequently forwarded messages across multiple groups, amplifying the spread of content with short but intense lifecycles (mean lifespan=6.8 days). Network analysis showed that users typically joined multiple groups in rapid succession and that conspiratorial messages traveled across groups within weeks. We underscore the importance of user-centric monitoring, time-sensitive interventions, and platform-specific models for content detection.

## 1. Introduction

Telegram has emerged as a key venue for digital discourse during crises, particularly in environments where users seek alternative narratives outside of mainstream media channels. Telegram provides fertile ground for the proliferation of misinformation, conspiracy theories, and other forms of agenda-driven content [15, 17]. During the COVID-19 pandemic, Telegram gained traction in Singapore as a platform for discussing health policies, personal experiences, and political opinions, often reflecting or reinforcing skepticism toward institutional authority [12].

Singapore offers a distinctive case for studying online misinformation. With one of the highest social media penetration rates globally and a government highly invested in managing information flow, it presents a media landscape intersecting with increasingly decentralized and unmoderated platforms like Telegram [16, 14, 1]. Previous work has shown how these online spaces can function as echo chambers for anti-vaccine sentiment, political discontent, and conspiracy theories, particularly during moments of public health uncertainty [3, 11, 5]. Yet there remains limited research on the structure and dynamics of these conversations within closed messaging systems in Southeast Asia [10]. This gap is critical given that Southeast Asia represents one of the most digitally connected yet politically diverse regions in the world. Platforms like Telegram are increasingly used not only for social mobilization but also for the dissemination of information that can influence public opinion, health behaviors, and political stability.

In this study, we analyzed Singapore-based Telegram groups. Our aim was to characterize the ecosystem of conspiratorial discourse in Telegram by asking: How is conspiratorial content disseminated within Singapore-based Telegram groups, and what roles do users play in shaping its spread, movement across groups, and message lifecycle? By mapping both user-level behaviors and message-level patterns, this study contributes to a deeper understanding of how conspiratorial narratives operate in low-moderation environments [13, 4]. These insights can inform both technical and policy strategies to mitigate the risks of misinformation and build more resilient digital information systems [9].

## 2. Methods

### Data collection and preparation

We first selected three content experts who had published at least ten peer-reviewed articles in the last three years around misinformation, and conspiracies, defined broadly. We ensured the content experts conducted research in the Singapore context. Content experts separately developed lists of Telegram groups most relevant to COVID-19 in Singapore. Groups were chosen based on the number of users in the group, how long the group had been active, and number of messages. Each expert developed a list of ten groups independently, and we selected only Telegram groups common to the three experts’ lists. The groups selected were: Healing the Divide Discussion, CovidSurvivors, SG Corona Freedom Lounge. Using tools within Telegram, we downloaded all chats in the groups since inception. Data from the groups was merged into a single file.

### Data processing and overview

Annotation for the conspiratorial content classifier was conducted by two content experts who had published more than five peer-reviewed papers in the broad misinformation space. Label agreement was 85%, and a third expert resolved any disagreements. We conformed to the definition of conspiratorial content established by past work [6]. The definition is as follows: A conspiracy theory is a set of narratives designed to accuse an agent(s) (be they individuals, groups, or organizations) of committing a specific action(s), which is believed to be working towards a secretive and malevolent objective(s) (secret plot). A total of 10000 messages were annotated, to create a balanced dataset of 1000 conspiratorial messages and 1000 non-conspiratorial messages. We tokenized, lemmatized, and normalized the corpus to prepare the texts for model training. We extracted several text features for model training: ngrams; word embeddings; TF-IDF. We used latent Dirichlet allocation (LDA) for topic modeling. Content experts decided the number of topics independently and then met to decide on the final topic model solution. Topic labels were assigned by GPT-4.

### Classification

Classification was performed using a range of machine learning algorithms, with an 80/20 train-test split, and data balanced 50:50 across classes. Both models were tested on the same data, 50:50 split between Telegram data (1=conspiratorial content; 0=non-conspiratorial content). We developed the following model: Classifier trained only on Telegram data (1=conspiratorial content; 0=non-conspiratorial content).

### Network analysis

We performed network analysis with igraph to detail movement to additional groups, differences in edge characteristics, for example, forwarded vs non-forwarded messages, among other analyses.

## 3. Results

Table 1 provided an overview of the Telegram groups. There were 19,696 unique users across all three groups. Topics were centered on pandemic skepticism, vaccine risks, and the questioning of mainstream medical narratives. Mean length of all messages was 25.7 words (SD=42.8). Word embedding analyses indicated that users linked the WEF (World Economic Forum) to globalist conspiracies and a new world order.

**Table 1.** Telegram Groups Examined for (Non) Conspiratorial Content.

| Dataset | Words | Messages | Unique Users | Dates (From - To) |
| --- | --- | --- | --- | --- |
| Healing the Divide Discussion | 8,672,597 | 366,183 | 13,950 | 2025-01-20 - 2021-08-11 |
| SG Corona Freedom Lounge | 1,130,351 | 66,016 | 4,558 | 2025-01-19 - 2021-06-14 |
| CovidSurvivors | 395,965 | 21,656 | 3,859 | 2025-01-19 - 2021-07-31 |

We presented metrics for the best performing classification model in Table 2. We used several classification algorithms and features, and found that the model using only TF-IDF performed reasonably well. LLMs (e.g., BERT, Llama) did not perform as well, perhaps due to limited data.

**Table 2.**
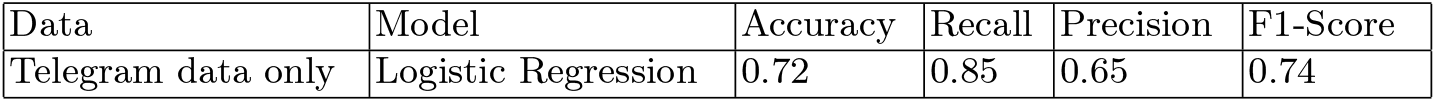
Model Metrics.

Telegram data only classifier was then used to classify all the text data in our study. We extracted a class-balanced sample of 200 items. Content experts verified accuracy of labels to be 72%. Example unigrams for the labelled data were as follows: Gates; pandemic; funded. Below are examples of text:

1. Bill Gates and Clinton foundations + big pharmas are pushing for mRNA vaccine worldwide because they want to condition the majority to be open to such a novel technique;
2. yup, there’s a theory that BG wants to use vaccines for population control in developing countries (ie make the population infertile);
3. Covid vaccines are BIOWEAPONS being used for genocide and depopulation;

Table 3 detailed that the bulk of users (90%) did not frequently post conspiratorial content. However, for 4.8% users, over 10% of their content was labelled as conspiratorial. Mean length of messages was 29.3 words (SD=51.2).

**Table 3.** Distribution of conspiratorial content by users.

| Percentage of conspiratorial content | N of users (% of total users) |
| --- | --- |
| 0% | 17654 (89.63%) |
| >0-10% | 1117 (5.67%) |
| 10-20% | 628 (3.18%) |
| >20% | 297 (1.50%) |

### Network analysis

#### Message patterns

Comparing forwarded vs nonforwarded messages (e.g., replies), we noticed that forwarded messages tended to refer to specific organizations and people, when describing various conspiracies. Nonforwarded messages tended to be less specific. Forwarded messages (M=25.9 words, SD=50.3) were slightly longer than nonforwarded messages (M=21.6, SD=38.5). Table 4 indicated that users who were in more groups tended to forward more messages. We suggest that users common across groups were more active participants.

**Table 4.** User characteristics by forwarding.

| N of groups | Mean (SD) of forwarded messages | % of conspiratorial content |
| --- | --- | --- |
| 1 | 15.7 (224.3) | 9.2 |
| 2 | 63.8 (376.3) | 10.2 |
| 3 | 147.6 (600.2) | 7.3 |

**User behaviors** Figure 1 indicated that users moving to additional groups spiked in the first two weeks of August. Most users who joined an additional group (n=288, %=13.8) seemed to join another group in the first 24 hours after joining a previous group. We suggest that users joined the available conspiratorial groups in relatively quick succession. There was limited movement to additional groups after 2021. Table 5 indicated the breakdown of users who were present in more than one group. 465 users were present in all three groups.

**Table 5.** Users present in additional groups.

|  | Healing the divide | SG Corona Freedom Lounge |
| --- | --- | --- |
| SG Corona Freedom Lounge | 1084 | - |
| CovidSurvivors | 1232 | 820 |

**Fig. 1.**
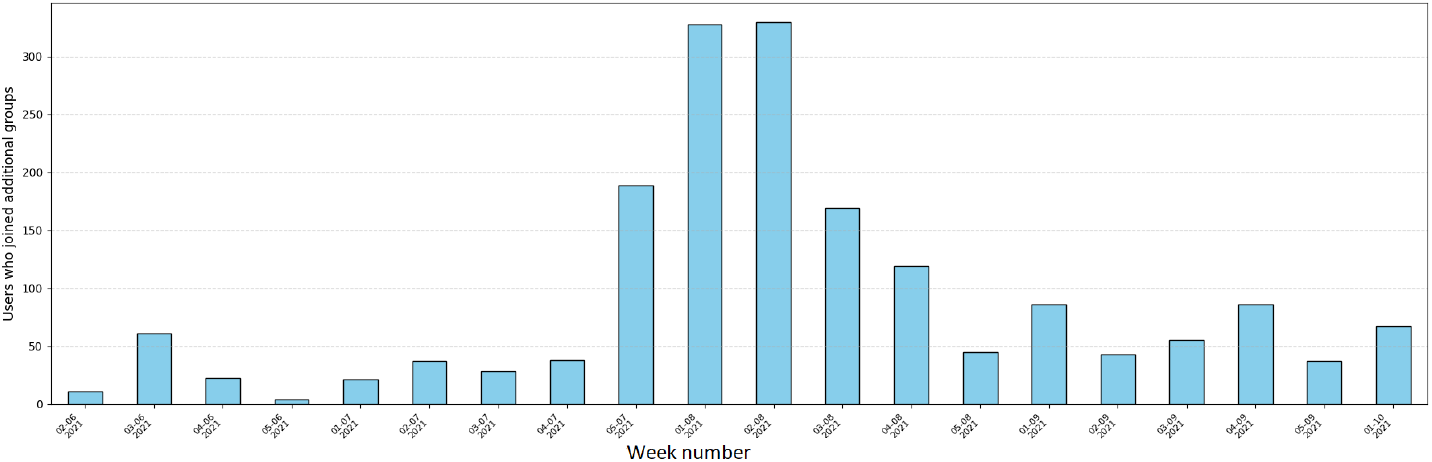
Dates of when users join an additional group

Figure 2 detailed movement of users (10+% conspiratorial content) to an additional Telegram group. Users from SG Corona Freedom Lounge seemed the most likely to join additional groups, with the bulk of them joining Healing the Divide Discussion. We suggest that certain groups attract users who tend to post conspiratorial content, and these users may seek to proliferate their content in other groups. This finding is in accordance with Figure 1 which detailed a spike in users moving to additional groups spiked.

**Fig. 2.**
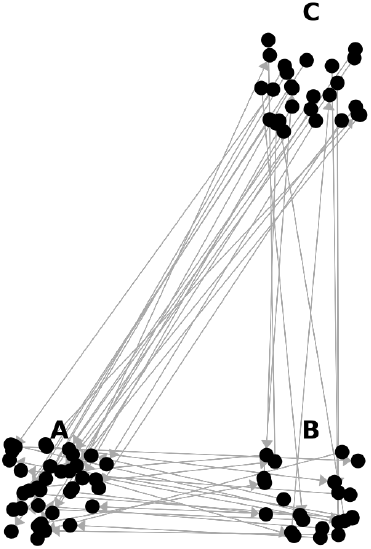
Users moving to an additional group. Node represents user, edge represents joining an additional group. A=Healing the Divide Discussion, B=CovidSurvivors, C=SG Corona Freedom Lounge

Figure 3 detailed the timeline of messages that were shared more than five times in our dataset. Each dot is when a message was shared, and a subsequent dot in the same line represents the next sharing of the message. Table 6 indicated a sample of the messages shared in Figure 3. We only included messages that were not generic messages sent by moderators. Messages broadly concerned vaccinerelated conspiracies. Messages were present in groups for roughly similar periods (M=6.8 days, SD=6.3). We suggest that conspiratorial content has a specific lifecycle in conspiratorial-aligned groups before it gets replaced by other content.

**Table 6.** Full messages.

| Message ID | Sentence |
| --- | --- |
| Message 1 | "When Lee Hsien Loong took over from the seat-warmer PM Goh Chok Thong, he was quite oblivious to the sacrifices made by civil servants like us to make it possible for him and his cabinet stooges to sit on their fat asses to enrich themselves with millions of people's money to wallow in the luxuries of their high office. |
| Message 2 | "We are now at a crossroads worldwide. The time is upon us to decide if we are going to allow this type of medical fascism to persist, and impact upon the futures of our children. Or if we are finally going to say no to tyrannical government policy." |
| Message 3 | Supreme Court ruling Hardly anyone noticed that Robert F. Kennedy Jr. won the case against all the pharmaceutical lobbyists. Covid vaccines are not vaccines. In the ruling, the Supreme Court confirms that the damage caused by Covid mRNA gene therapies is irreparable. |
| Message 4 | I just saw this message from some concerned member here, not sure if the new noravax so safe after all |
| Message 5 | The TRAITORS in the UK government admits that vaccines have damaged the natural immune system of those who have been double-vaccinated. |
| Message 6 | "We will contact you for the proof of your termination/unemployment status / unvaxxed status for you to attend it on the scholarship. Proceeds from this workshop will go towards paying for the lawyer for this session with extra going towards our legal actions against MOH for defamation." |

**Fig. 3.**
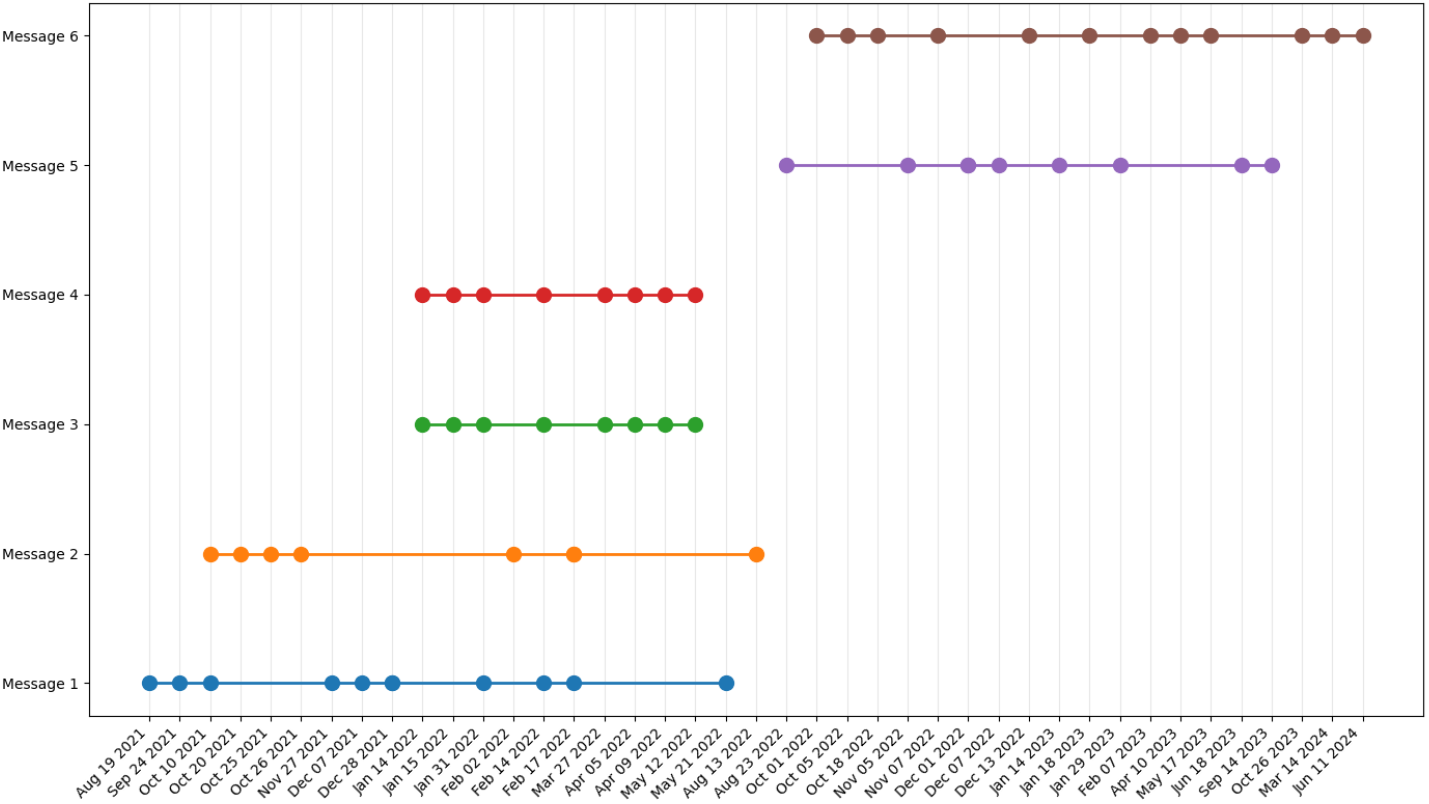
Movement of phrases with time in healing group

Figure 4 detailed the movement of messages in Table 6 across groups. Each node denoted when a message was shared by an additional user. Messages were shared an average of 4.3 times (SD=4.6) before they moved to an additional group. Messages took an average of 0.25 months (SD=0.5) to move from one group to another. We suggest that conspiratorial content has a limited lifecycle, possibly dependent on the group it circulates in.

**Fig. 4.**
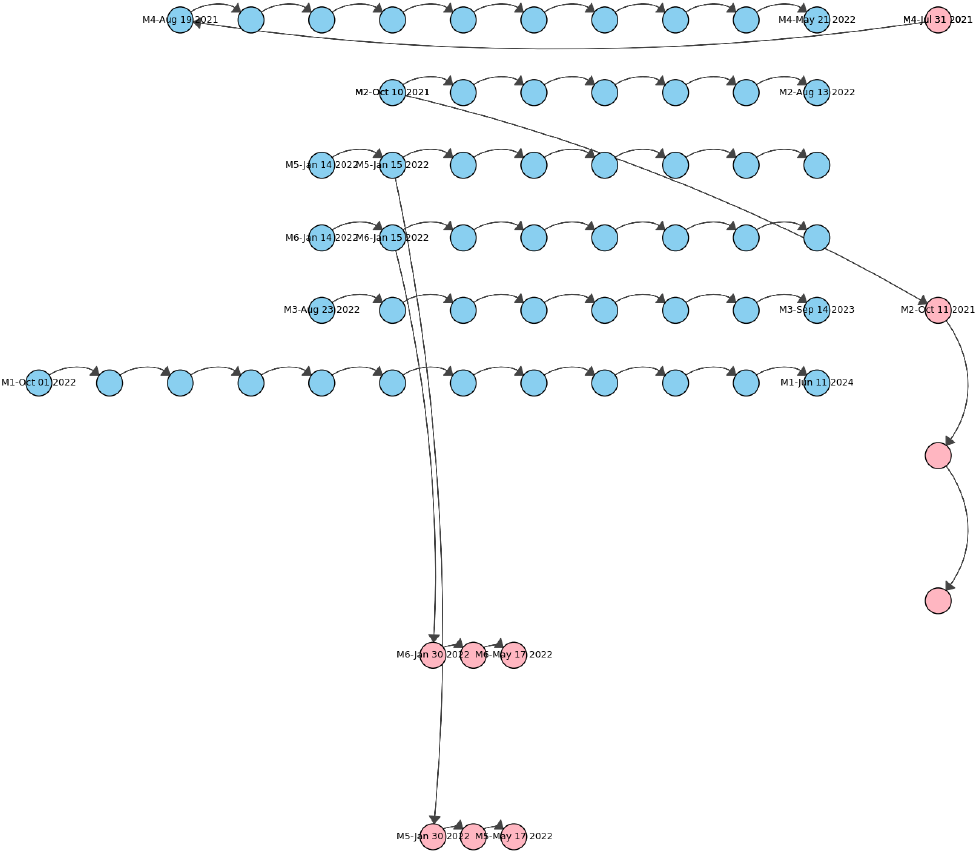
Blue nodes: Group Healing the Divide; Pink Nodes: SG Corona Freedom Lounge. Numbers beside nodes refer to the messages in Table 6. Only two groups had messages that were shared across groups. Dates: Timestamp for message lifecycle start, end, or movement to an additional group.

## 4. Discussion

Several key findings emerged. First, although a large portion of users (approximately 90%) did not frequently post conspiratorial content, a small yet vocal minority (under 5%) may be responsible for a disproportionately high volume of such material. Thematically, conspiratorial content focused on familiar tropes: elite control (e.g., Bill Gates, Klaus Schwab), geopolitical manipulation, vaccine danger, and depopulation agendas. Our topic models confirmed prior hypotheses about the persistence of such frames in online misinformation ecosystems. Word embedding analyses linked the World Economic Forum (WEF) to a “globalist agenda” and “new world order,” further emphasizing how international institutions were reframed as antagonistic actors. The lifecycle of conspiratorial messages was also notable. These messages tended to be shared within narrow windows of activity (mean=6.8 days), suggesting temporal bursts of attention rather than sustained discussions. Movement across groups was infrequent but rapid, typically occurring within 24 hours. This pattern suggested that conspiratorial information functioned much like a viral packet, transmitted quickly to receptive audiences before fading or being replaced.

While the classification model achieved strong in-sample performance, its reliability depends on the characteristics of the training data [8, 2]. Although expert validation provided a measure of accuracy, future applications may require retraining or recalibration to account for linguistic drift or shifts in narrative style over time [7].

## Data Availability

All data produced is available on request

